# Comparison of COVID-19 Antigen Rapid Test (Oral Fluid) and Real-Time RT-PCR in the laboratory diagnosis of SARS-CoV-2 infection

**DOI:** 10.1101/2022.10.07.22280607

**Authors:** Lotte M. Mense, Sander Ouburg

**Affiliations:** Microbe&Lab BV, dept. Research & Development, Amsterdam, the Netherlands

## Abstract

Severe acute respiratory syndrome coronavirus-2 (SARS-CoV-2) was first diagnosed in December 2019. Since then this virus has caused an ongoing wide pandemic. Accurate diagnostic tests for SARS-CoV-2 are used to prevent the virus from spreading. However, these tests could not keep up with the demand and were not available in all places. Self-testing devices are easy-to-use-tests and reduce the demand in the diagnostic laboratories. The Antigen Rapid Test evaluated in this study uses oral fluid which is a non-invasive technique compared to nasopharyngeal swabs.

In this study the COVID-19 Antigen Rapid Test (Oral fluid) was evaluated with 150 SARS-CoV-2 positive saliva specimens and 350 SARS-CoV-2 negative saliva specimens. The Antigen Rapid Test was performed according to the instruction manual. SARS-CoV-2 Real-time RT-PCR was used as Golden Standard.

Although the criteria of the WHO are specific to nasal / nasopharyngeal samples (and not saliva), the specificity of the Antigen Rapid Test meets the criteria of the World Health Organization (WHO; specificity ≥ 97%). The test meets the WHO sensitivity criteria in samples with higher viral loads (Ct<30), showing the better performance of the test in highly positive samples. For positive SARS-CoV-2 specimens with a Ct value lower than 30 a sensitivity of 83.8% (95% CI: 80.1%-86.8%) and an accuracy of 95.9% (95% CI: 93.7%-97.4%) was observed. This shows that this assay with saliva samples is able to meet the high standards set by the WHO. The performance of the test is comparable to other antigen rapid tests reported in meta-analyses. Furthermore, the test allows self-testing which is non-invasive, affordable and straightforward. This antigen rapid test may provide an affordable, quick, and easy to perform method to differentiate between individuals with high and low viral loads.

## Introduction

In December 2019, the first humans with respiratory signs and symptoms in the world were diagnosed with a novel coronavirus (Simmons & Schur, 2022). This virus was initially named novel coronavirus-2019 (nCOV-19) by the World Health Organization (WHO). Later the virus was renamed as severe acute respiratory syndrome coronavirus-2 (SARS-CoV-2) (Niaz & Nisar, 2021). The virus has infected humans and animals rapidly and this has caused an ongoing wide pandemic.

In many countries accurate diagnostic tests for SARS-CoV-2 became broadly available to prevent the virus from spreading rapidly. Unfortunately, in many places either accurate diagnostic tests were not available or the laboratories could not keep up with the demand (World Health Organization, 2020). In these places point of care tests, such as Antigen Rapid Tests were used. These self-testing devices are rapid and easy-to-use tests. Over time these tests have become more acceptable since it reduces the demand in the diagnostic laboratories for SARS-CoV-2 (World Health Organization, 9 March 2022).

Currently there are two types of SARS-CoV-2 Antigen Rapid Tests. One uses nasal swabs and the other uses oral fluid. Nasal swabs are considered as the Golden Standard. However, using saliva as specimen for viral infection diagnostics has gained more interest. Collecting saliva specimens is a non-invasive technique, economical and straightforward. Studies have shown that SARS-CoV-2 Antigen Rapid Tests based on saliva is a valid alternative for nasopharyngeal swabs (Basso, et al., June 2021).

In this study the performance of COVID-19 Antigen Rapid Test (Oral fluid) from Hangzhou Alltest Biotech was analysed. The Antigen Rapid Tests is a membrane-based immunoassay for the detection of SARS-CoV-2 nucleocapsid protein antigens in human oral fluid specimens. The device was evaluated using 500 patient saliva samples. Of these 500 samples, 150 samples were SARS-CoV-2 positive and 350 samples were SARS-CoV-2 negative. The PerkinElmer SARS-CoV-2 RNA isolation kit & SARS-CoV-2 Real-time RT-PCR assay was used as gold standard.

## Methods

### Specimen collection

For this study 500 saliva specimens were used, 350 SARS-CoV-2 negative specimen and 150 SARS-CoV-2 positive specimen. Before collecting oral fluid, the patients were not allowed to place anything in the mouth including food, drink, gum, or tobacco products for at least 10 minutes prior to collection. The saliva specimens were self-collected by patients themselves by spitting into sterile tubes. Samples were submitted for routine SARS-CoV-2 screening and were not from a high risk or highly symptomatic population. The rest materials were used anonymously for this evaluation.

### Method SARS-CoV-2 Antigen Rapid Test (Oral Fluid)

This study evaluates the COVID-19 Antigen Rapid Test (Oral Fluid). This lateral flow SARS-CoV-2 Antigen Rapid Test was performed according to the instruction manual. In short: 500μL specimen was collected and buffer was added to the specimen. Then 2 drops were added on the sample well of the cassette. The results were read at 15 minutes.

### Method DNA/RNA isolation and PCR

DNA/RNA isolation and PCR was performed according to the instructions for SARS-CoV-2 RT-qPCR Reagent kit of PerkinElmer. In short,⍰a mixture of⍰300μl⍰containing⍰lysis buffer, ⍰poly A, and proteinase K⍰(75:1:2.5)⍰ was added to⍰300μl of⍰specimen. The default⍰Chemagic⍰Viral300 360 H96 prefilling short VD200406 protocol on the⍰Chemagic™⍰360 2040-0020 (PerkinElmer) was used for the isolation. An elution volume of 100μl⍰was chosen.⍰

The PCR targets in this assay are the SARS Cov-2 N-gene⍰, the SARS Cov-2 ORF1ab-gene,⍰and⍰the⍰human⍰leukocyte⍰antigen (HLA-)gene (internal⍰positive⍰control (IPC)). The PCR master mix contains per sample, 1μL⍰CoV2 Reagent A,⍰and⍰5μL⍰CoV2 Enzyme Mix (SARS-CoV-2 RT-qPCR Reagent kit, PerkinElmer). 5μL PCR reagent mix and 10μL extracted nucleic acid was used per reaction. The⍰QuantStudio™ 5 Real-Time PCR System (ThermoFisher⍰Scientific Applied Biosystems) was used for running rt-RT-qPCRs.

⍰A negative control (demi water) and an independent run control (IRC, previously tested positive sample) were taken along for each isolation.⍰IRCs were created by pooling positive specimens until 5ml is obtained and diluting this in 45ml PBS.⍰For⍰each⍰PCR run, a⍰positive control (PC) was included.⍰Samples with clear S-curves and a Ct-value for the IPC were considered valid. Of these valid samples, samples with⍰a Ct-value for⍰either one or⍰both SARS-CoV-2 target genes (N and ORF1ab) were considered positive as per manufacturer’s instructions. The whole procedure was repeated for samples with invalid results.

## Results

### Valid results were obtained for all samples with the PCR and the antigen rapid test

Excluding positive SARS-CoV-2 specimens with a Ct value higher than 30 from the calculations, resulted in a higher sensitivity and an accuracy for the Antigen Rapid Test. Sensitivity increased to 83.8% (95% CI: 80.1%-86.8%) and accuracy increased to 95.9% (95% CI: 93.7%-97.4%). Results are shown in tables 1 and 2.

**Table 1:** Results COVID-19 Rapid Antigen Test was detected in different CT values of positive specimens.

| CT Value | Rapid Test Positive | RT-PCR Positive | Sensitivity(%) |
| --- | --- | --- | --- |
| CT<25 | 70 | 71 | 98.6%(95% CI: 96.9%-99.4%) |
| CT<30 | 98 | 117 | 83.8%(95% CI: 80.1%-86.8%) |
| Ct<37 | 100 | 150 | 66.7%(95% CI: 62.4%-70.7%) |

**Table 2:** Results COVID-19 Rapid Antigen Test of negative specimens.

| Rapid Test negative | RT-PCR Negative | Specificity(%) |
| --- | --- | --- |
| 350 | 350 | 100%(95% CI: 99.1%-100%) |

## Discussion

The specificity in this experiment was comparable to the specificity described in the instruction manual of the COVID-19 Antigen Rapid Test. The instructions for use describe a sensitivity of 90.1% (95% CI: 82.5%-95.1%), a specificity of 99.3% (95% CI: 97.7-99.9%), and an accuracy of 97.0% (95% CI: 94.9%-98.5%). The study described in the IFU was performed with 101 SARS-CoV-2 positive and 305 SARS-CoV-2 negative specimens. Differences in the studied population and viral load may contribute to the differences in the study outcome.

Antigen Rapid Tests provide an economical and easy-to-perform platform for SARS-CoV-2 testing. Sample preparation can usually be done in less than five minutes and results can usually be read after fifteen minutes, making the total time to result less than half an hour. The assays are small, require little or no equipment besides materials provided in the kit, and can usually be performed on location. NAAT tests often need to be performed on a more specialised location by qualified personnel. Sample preparation for NAAT tests and the running of the NAAT assay may take up to two hours. Saliva samples are less invasive for the patient to obtain as it does not require a nasal, nasopharyngeal, or oropharyngeal swab. The study of Basso et al. showed saliva to be a valid alternative to a nasopharyngeal swab in early infections (Basso, et al., June 2021).

In 2020 the World Health Organization published guidelines for point of care tests for suspected SARS-CoV-2 cases and their close contacts (World Health Organization, 2020). The Antigen Rapid Tests were used in areas in which there was a lack of technical expertise and inadequate laboratory capacity, RT-PCR testing not broadly available, or diagnostics could not keep up with the demand. The criteria of the WHO state that the limit of detection should be a nasal or nasopharyngeal swab specimen with a Ct value between 25 and 30. The sensitivity should be ≥ 80%, but preferably ≥90%. The specificity should be ≥ 97% and preferably >99% (World Health Organization, 2020). The Antigen Rapid Test was capable of detecting SARS-CoV-2 in the saliva samples, although with a lower sensitivity compared to RT-PCR (PerkinElmer). A negative result in the antigen rapid test does not exclude a SARS-CoV-2 infection and verification of negative results by PCR would be advised.

The higher the RT-PCR CT value is, the lower virus load the sample contains, which may be harder for the Antigen Rapid Test detection.

As the results in table 1 show highest sensitivity was observed in samples with a Ct value below 25 (98.6%). Sensitivity decreases to 83.8% in samples with a Ct value below 30 and keep decreased to 66.7% in samples with a Ct value below 37. Therefore we can conclude that the Antigen Rapid test performs better with samples with a high viral load. Performance of the COVID-19 Antigen Rapid Test is comparable to other antigen rapid tests reported in meta-analyses (Brümmer, et al., 2021) (Cochrane Infectious Diseases Group, et al., 2020), which also show that antigen rapid tests perform better in symptomatic individuals with higher viral loads.

In the last two years interest in Antigen Rapid Tests has grown since users can reliably and accurately self-test. Self-testing has become more acceptable and feasible, and may reduce the burden on existing diagnostics of SARS-CoV-2. Furthermore self-testing combined with self-quarantine may help reduce spread of SARS-CoV-2 (Brümmer, et al., 2022). The guidelines of the WHO state a sensitivity of ≥ 80% and a specificity of ≥ 97% for antigen rapid tests using nasal and/or naso-pharyngeal samples, but these criteria are for individuals with symptoms of SARS-CoV-2 (World Health Organization, 9 March 2022). The results of this study show that the COVID-19 Antigen Rapid Test with saliva samples is able to provide sensitivity and specificity in high viral load samples that meets the standards set by the WHO for SARS-CoV-2 antigen rapid tests. In this study it is unknown whether the patients had symptoms during collecting the specimens. However, a low Ct value, i.e. a high viral load, is more common when people have the most symptoms. The Ct value increases over time when the SARS-CoV-2 infection is resolved by the immune system. Assuming that the specimens with a Ct value lower than 30 were collected from mostly symptomatic individuals, the Antigen Rapid Test has a high sensitivity. This test may offer a quick and easy to perform method to identify individuals with a high viral load who may be more contagious compared to individuals with a low viral load.

## Conclusion

In summary, COVID-19 Antigen Rapid Test Oral fluid allows self-testing which is non-invasive, economical and straightforward. The study result of SARS-CoV-2 positive specimens with a Ct value lower than 25 showed a higher sensitivity of 98.6% (95% CI: 96.9%-99.4%) and an accuracy of 99.8% (95% CI: 98.7%-100%). The study result of SARS-CoV-2 positive specimens with a Ct value lower than 30 showed a sensitivity of 83.8% (95% CI: 80.1%-86.8%) and accuracy of 95.9% (95% CI: 93.7%-97.4%). The overall sensitivity of the Antigen Rapid Tests was 66.7% and the specificity was 100% in samples with a CT value lower than 37. The Antigen Rapid Test performed better in specimens with high viral loads. This antigen rapid test may provide an affordable, quick, and easy to perform method to differentiate between individuals with high and low viral loads.

## Supporting information

Conflict of interest form

## Data Availability

All data produced in the present study are available upon reasonable request to the authors

## References

Basso, D., Aita, A., Padoan, A., Cosma, C., Navaglia, F., Moz, S., Plebani, M. (June 2021). Salivary SARS-CoV-2 antigen rapid detection: A prospective cohort study. Clinica Chimica Acta, volume 517 pages 54-59.

Brümmer, L. E., Erdmann, C., Tolle, H., McGrath, S., Olaru, I. D., Katzenschlager, S., Denkinger, C. M. (2022). The clinical utility and epidemiological impact of self-testing for SARS-CoV-2 using antigen detecting diagnostics: a systematic review and meta-analysis. MedRxiv.

Brümmer, L. E., Katzenschlager, S., Gaeddert, M., Erdmann, C., Schmitz, S., Bota, M., Denkinger, C. M. (2021). Accuracy of novel antigen rapid diagnostics for SARS-CoV-2: A living systematic review and meta-analysis. PLOS Medicine, e1003735.

Cochrane Infectious Diseases Group, Dinnes, J., Deeks, J. J., Adriano, A., Berhane, S., Davenport, C.,. Cochrane COVID-19 Diagnostic Test Accuracy Group. (2020). Rapid, point-of-care antigen and molecular-based tests for diagnosis of SARS-CoV-2 infection. Cochrane Database Syst. Rev, CD013705.

Niaz, K., & Nisar, M. F. (2021). Coronavirus Disease-19 (COVID-19): A Perspective of New Scenario. Singapore: Bentham Science Publishers.

Simmons, D. P., & Schur, P. H. (2022). Detection of SARS-CoV-2 Antibodies in Diagnosis and Treatment of COVID-19, An Issue of the Clinics in Laboratory Medicine. Philadelphia: Elsevier Health Sciences.

World Health Organization (2020). Target product profiles for priority diagnostics to support response to the COVID-19 pandemic v.1.0. Geneva, Switzerland.

World Health Organization. (9 March 2022). Use of SARS-CoV-2 antigen-detection rapid diagnostic tests for COVID-19 self-testing.

